# Association between sleep apnoea and arrhythmia: A multivariable Mendelian randomization study

**DOI:** 10.1101/2025.04.02.25325145

**Authors:** Wu He, Jian Wang, Jiali Nie, Dao Wen Wang, Li Ni, Dan Li

## Abstract

**Background:** Sleep apnoea is recognized as a cardiovascular risk factor, especially in individuals with existing heart conditions. Understanding the direct causal relationship between sleep apnoea and arrhythmia, while accounting for confounding cardiovascular factors such as coronary artery disease, hypertension, heart failure, and myocardial infarction, is essential for effective cardiovascular risk management.

**Methods:** Utilizing data from the FinnGEN study spanning 1998 to 2019, we conducted a Mendelian randomization analysis on a European cohort to explore the association between genetically predicted sleep apnoea and various arrhythmias, including atrioventricular block, atrial fibrillation, and ventricular arrhythmia. The study aimed to identify specific genetic components contributing to these associations.

**Results:** The genetic analysis demonstrated significant associations between a genetic predisposition to sleep apnoea and certain arrhythmias. Importantly, sleep apnoea showed an independent association with atrioventricular block, even after adjusting for other cardiovascular conditions. Two single nucleotide polymorphisms, rs9937053 and rs10507084, were identified as critical factors in the causal pathway connecting sleep apnoea to atrioventricular block. However, there were no direct causal relationships found between sleep apnoea and the other arrhythmias examined.

**Conclusions:** This study emphasizes the critical role of considering sleep apnoea in cardiovascular management, particularly due to its direct causal link to atrioventricular block influenced by specific genetic variants. These findings call for additional research to further explore the impact of sleep apnoea on other types of arrhythmias.

**What Is New?:**

- This study establishes a direct causal link between sleep apnoea and atrioventricular block using Mendelian randomization in a large European cohort from the FinnGEN study.
- Two genetic markers, rs9937053 and rs10507084, are identified as critical components in the causal pathway connecting sleep apnoea to atrioventricular block, highlighting potential targets for future intervention.
- The study also uncovers bidirectional causal relationships between sleep apnoea and hypertension, and sleep apnoea and obesity-related traits like BMI, suggesting complex interactions that affect cardiovascular and metabolic health.

**What Are the Clinical Implications?:**

- The findings emphasize the importance of considering sleep apnoea in cardiovascular management, particularly concerning its direct causal role in atrioventricular block influenced by specific genetic variants.
- Including sleep apnoea in the clinical assessment and management strategies for patients with or at risk of atrioventricular block could enhance individualized patient care and improve cardiovascular outcomes.
- The identification of specific genetic variants associated with sleep apnoea-related arrhythmias suggests new avenues for research and the development of targeted therapies, potentially leading to more effective prevention and treatment approaches for cardiovascular comorbidities in patients with sleep apnoea.

## Introduction

Sleep Apnoea (SA), comprising Obstructive Sleep Apnoea (OSA), Central Sleep Apnoea (CSA), and Cheyne-Stokes Breathing (CSB), represents a pervasive health issue affecting over 1 billion adults globally^1^. The physiological stressors induced by SA exert enduring biological effects, leading to cardiovascular structural alterations that heighten the risk of cardiac arrhythmogenesis^2^. Cardiac arrhythmias, whether occurring independently or in conjunction with cardiovascular disorders, are commonly observed in structural heart diseases like coronary artery disease, cardiomyopathy, and myocarditis, particularly in the context of heart failure or acute myocardial infarction^3^. A notable prevalence of SA and arrhythmias is evident among individuals with these cardiac conditions, with epidemiological evidence demonstrating a significant coexistence of these comorbidities^2^.

Community-based studies consistently reveal a strong correlation between Sleep-Disordered Breathing (SDB) and nocturnal cardiac arrhythmias. Individuals with moderate to severe SDB are nearly five times more likely to develop atrial fibrillation (AF) compared to those without SDB, even after adjusting for factors such as obesity and pre-existing cardiovascular risk^4^. OSA, but not CSA, has been shown to alter the balance between the sympathetic and parasympathetic nervous systems, as indicated by heart rate variability measures, contributing to a higher incidence of AF over an average follow-up period of 8.0±2.6 years^5^. Retrospective analyses ranging from 2 to 11 years reveal an independent association between the onset of AF and the severity of OSA, as well as its diagnosis^6,7^. In contrast, prospective cohort studies suggest that CSA, rather than OSA, is linked to AF^8^. Furthermore, severe SDB is independently associated with an increased risk of Non-Sustained Ventricular Tachycardia (NSVT) in both cross-sectional and prospective analyses^4,9^.

Research on Mendelian Randomization (MR) is increasingly focusing on the causal relationship between sleep apnoea and atrial fibrillation^10,11^. However, there remains a significant gap in comprehensive, multifactorial MR studies exploring sleep apnoea and various other types of cardiac arrhythmias. Against this backdrop, multivariable two-sample MR demonstrates clear advantages. It effectively controls for confounding factors through genetic variation, eliminating interference from reverse causation^12^. By analyzing multiple related factors, it enhances the accuracy of causal inferences. Additionally, by integrating data from two sample sets, it boosts statistical power and result robustness.

In this study, we applied a multivariable two-sample MR approach, using summary-level data from genome-wide association studies (GWAS), to establish a causal connection between sleep apnoea and specific subtypes of arrhythmia. The findings aim to inform future therapeutic strategies to mitigate these adverse cardiovascular outcomes.

## Materials and Methods

### Study assumption

To effectively apply MR, three critical assumptions must be satisfied^13^. First, the instrumental variable (IV), typically a genetic variant, must have a strong association with the exposure variables, such as sleep apnoea, coronary artery disease, hypertension, heart failure, and myocardial infarction. Second, the IV must be free from confounding with any unmeasured factors and should not directly affect the outcomes, including atrioventricular block, ventricular arrhythmia, atrial fibrillation, and supraventricular tachycardia. Lastly, the IV should influence the outcomes exclusively through its relationship with the exposure, without any alternative pathways.

### Exposure and data sources

We investigated a range of exposures, including sleep apnoea (Dataset: finn-b-G6_SLEEPAPNO), coronary artery disease (Dataset: ebi-a-GCST005195), hypertension (Dataset: ebi-a-GCST90038604), heart failure (Dataset: ebi-a-GCST009541), and myocardial infarction (Dataset: finn-b-I9_MI_STRICT). Comprehensive details are provided in the Supplementary Table 1. To pinpoint genetic variants linked to these conditions, we undertook an extensive search for pertinent GWAS (Genome-Wide Association Studies) through the MRC-IEU GWAS Catalog (https://gwas.mrcieu.ac.uk/). We prioritized GWAS studies with the largest participant cohorts for each exposure.

### Arrhythmia subtype and data sources

For the outcomes, we sourced summary-level data from a variety of research consortia. Specifically, for atrioventricular block, atrial fibrillation, ventricular arrhythmia, and supraventricular tachycardia, we utilized datasets from finn-b-I9_AVBLOCK (with 2,388 cases), ebi-a-GCST006414 (with 60,620 cases), ebi-a-GCST90018940 (with 1,018 cases), and ukb-b-11748 (with 1,306 cases), respectively. The data for supraventricular tachycardia were provided by the MRC-IEU Consortium. Further details regarding these datasets are outlined in the Supplementary Table 1.

### Statistical analysis

We extracted genetic variants (SNP, single nucleotide polymorphisms) strongly associated with exposure, defined as P < 5×10^−8^, and that were independent of one another, defined as a pairwise R^2^ < 0.001 based on the 10000 Genomes Project European ancestry superpopulation. Prior to MR analyses, we harmonized the files to ensure that the effect estimate of a given SNP was oriented to the same allele in all files. The causal effects of each exposure on outcomes were analyzed using the inverse-variance weighted (IVW) method, which provides a combined estimate of the causal effect from each SNP. IVW is equivalent to a two-stage least squares or allele score analysis using individual-level data and is commonly regarded as the conventional MR approach.

Associations between exposure and outcome were harmonized to ensure that estimates were aligned on the same allele. Ambiguous genetic variants with palindromic genotypes were excluded. We used the IVW method as the primary analysis, which combines SNP-specific estimates calculated by Wald ratios through dividing the genetic association with outcome by the genetic association with each exposure. Univariable and multivariable MR Analysis were employed. we scaled the odds ratios (ORs) and corresponding 95% confidence intervals (CIs) for outcomes to represent a one-standard deviation increase in exposure. In our MR analyses, the median weighted, MBE, and MR-Egger methods were considered as sensitivity analyses when investigating multiple genetic variants. Power calculations for MR were performed using the mRnd website (http://cnsgenomics.com/shiny/mRnd/). All analyses were conducted using R, version 4.2.0, for statistical computing. Specifically, the multivariable Mendelian Randomization analysis was performed using the ‘MendelianRandomization’ package, version 0.7.0, which facilitated the exploration of causal relationships between genetic variants and multiple phenotypes. The ‘TwoSampleMR’ package, version 0.5.6, was utilized for conducting two-sample MR analyses, and the ‘metaphor’ package, version 3.4.0, was applied for implementing multiple testing corrections. Custom scripts and functions developed for this study were also employed in conjunction with these packages to execute the complex analytical pipeline. The threshold for statistical significance was set at *P*≤0.0005(0.05/100) after Bonferroni correction. Associations with *P*≤0.05 but above the Bonferroni corrected significance threshold were considered suggestive of potential evidence for an association.

## Results

### Univariate MR Analysis on Sleep Apnoea, cardiovascular comorbidies and Arrhythmia

In a single-variable conventional MR analysis, we explored the causal relationships between sleep apnoea and various cardiovascular comorbidities—such as coronary artery disease, hypertension, myocardial infarction, and heart failure—and the risk of arrhythmia. The results indicated that both sleep apnoea (beta=0.347, *P*=0.045) and hypertension (beta=1.042, *P*=0.001), when examined separately, showed causal effect estimates associated with an increased risk of atrioventricular block, as detailed in Table 1. In contrast, coronary artery disease, myocardial infarction, and heart failure did not display causal effect estimates indicative of a higher risk of atrioventricular block (Table 1).

**Table 1.**
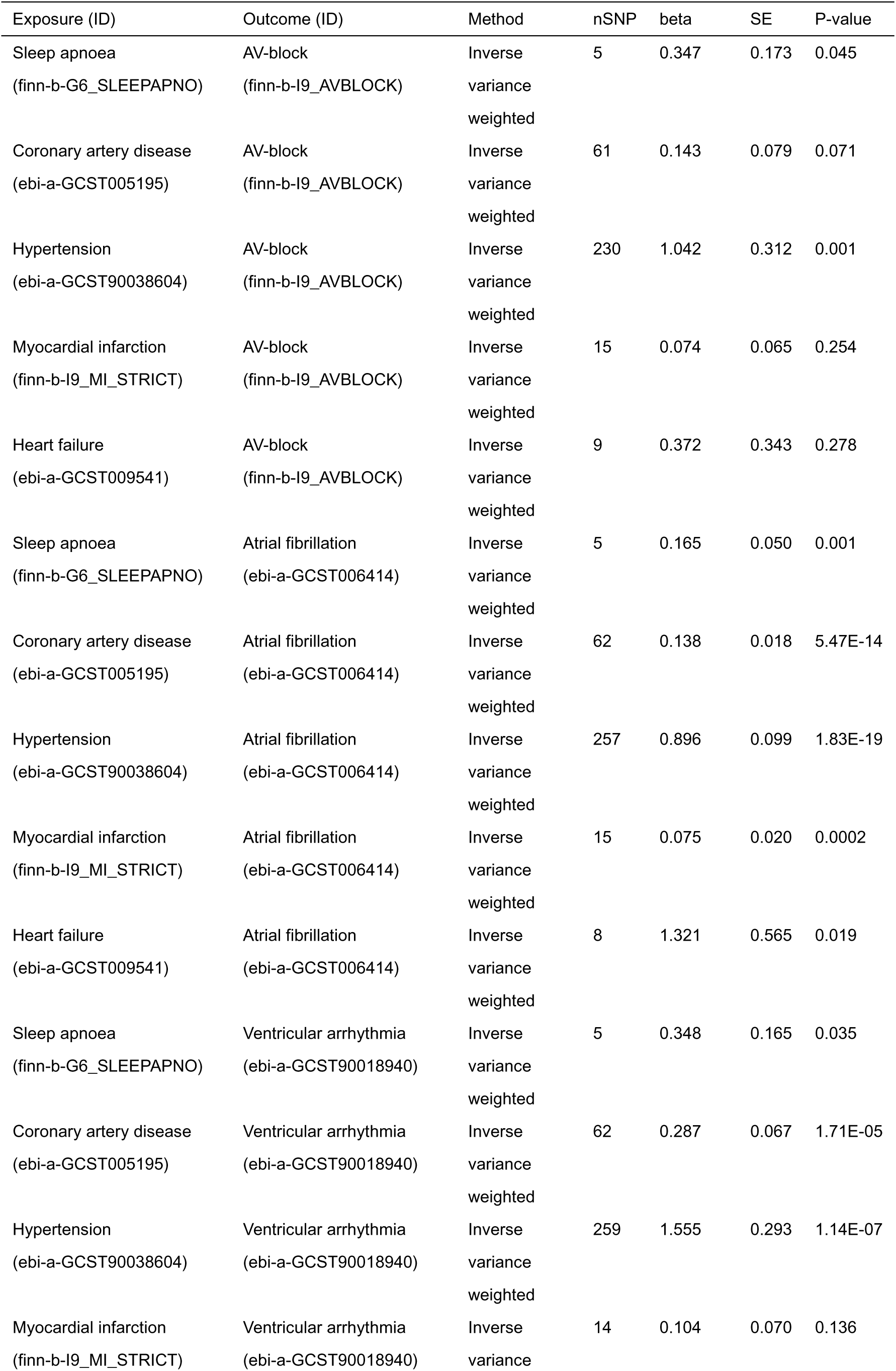
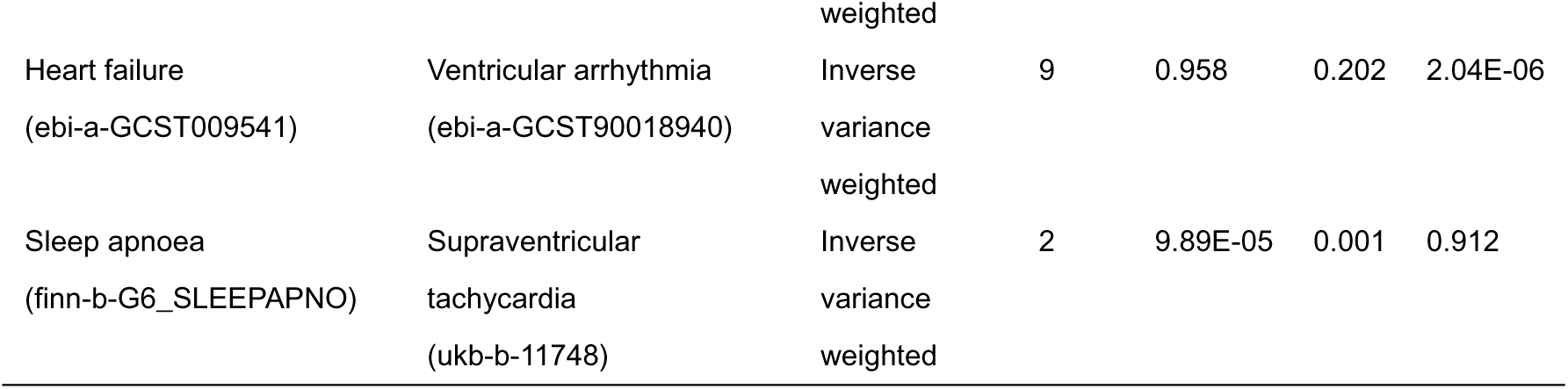
The causal effect of sleep apnoea or common cardiovascular related comorbidities with arrhythmia in the univariate Mendelian randomization analysis.

Univariate MR analysis further revealed a causal link between sleep apnoea or any of the examined cardiovascular comorbidities and atrial fibrillation, as shown in Table 1. Specifically, conditions such as sleep apnoea (beta=0.165, *P*=0.001), coronary artery disease (beta=0.138, *P*=5.47E-14), hypertension (beta=0.896, P=1.83E-19), myocardial infarction (beta=0.075, P=0.0002), and heart failure (beta=1.321, *P*=0.019) were all associated with an increased risk of atrial fibrillation (Table 1).

Additionally, univariate MR analysis revealed that, except for myocardial infarction, which does not have a demonstrated causal relationship with ventricular arrhythmia, sleep apnoea and other cardiovascular comorbidities examined in the study were found to have a causal relationship with ventricular arrhythmia (Table 1). The conditions associated with this increased risk include sleep apnoea (beta=0.348, *P*=0.035), coronary artery disease (beta=0.287, *P*=1.71E-05), hypertension (beta=1.555, *P*=1.14E-07), and heart failure (beta=0.958, *P*=2.04E-06) (Table 1). Lastly, the analysis indicated that sleep apnoea does not exhibit a causal relationship with supraventricular arrhythmia (Table 1).

### Multivariate MR Analysis: Sleep Apnoea and cardiovascular comorbidies on Arrhythmia

In the multivariable Mendelian randomization (MVMR) analysis, sleep apnoea maintained a strong causal relationship with atrioventricular block (OR=1.393, *P*=0.011), but showed no causal effect on atrial fibrillation (OR=1.021, *P*=0.765). In contrast, myocardial infarction (OR=0.886, *P*=0.040) and heart failure (OR=3.060, *P*=9.20E-24) were found to have causal effects on atrial fibrillation. Among the conditions studied, only heart failure exhibited a robust causal effect on ventricular arrhythmia (OR=1.497, *P*=0.036), as detailed in Table 2.

**Table 2.** Multivariable Mendelian randomization analysis of sleep apnoea and common cardiovascular comorbidities associated with arrhythmia.

| Exposure (ID) | Outcome (ID) | nSNP | beta | SE | P-value | OR | 95%CI |
| --- | --- | --- | --- | --- | --- | --- | --- |
| Sleep apnoea<br>(finn-b-G6_SLEEPAPNO) | AV-block<br>(finn-b-I9_AVBLOCK) | 2 | 0.331 | 0.130 | 0.011 | 1.393 | (1.080, 1.797) |
| Hypertension<br>(ebi-a-GCST90038604) | AV-block<br>(finn-b-I9_AVBLOCK) | 188 | 0.570 | 0.355 | 0.108 | 1.768 | (0.882, 3.543) |
| Sleep apnoea<br>(finn-b-G6_SLEEPAPNO) | Atrial fibrillation<br>(ebi-a-GCST006414) | 3 | 0.021 | 0.070 | 0.765 | 1.021 | (0.890, 1.172) |
| Coronary artery disease<br>(ebi-a-GCST005195) | Atrial fibrillation<br>(ebi-a-GCST006414) | 39 | -0.049 | 0.085 | 0.566 | 0.952 | (0.806, 1.125) |
| Hypertension<br>(ebi-a-GCST90038604) | Atrial fibrillation<br>(ebi-a-GCST006414) | 183 | -0.023 | 0.216 | 0.915 | 0.977 | (0.640, 1.192) |
| Heart failure<br>(ebi-a-GCST009541) | Atrial fibrillation<br>(ebi-a-GCST006414) | 5 | 1.118 | 0.111 | 9.20E-24 | 3.060 | (2.460, 3.806) |
| Myocardial infarction<br>(finn-b-I9_MI_STRICT) | Atrial fibrillation<br>(ebi-a-GCST006414) | 10 | -0.121 | 0.059 | 0.040 | 0.886 | (0.790, 0.995) |
| Sleep apnoea<br>(finn-b-G6_SLEEPAPNO) | Ventricular arrhythmia<br>(ebi-a-GCST90018940) | 3 | 0.175 | 0.122 | 0.152 | 1.191 | (0.938, 1.513) |
| Coronary artery disease<br>(ebi-a-GCST005195) | Ventricular arrhythmia<br>(ebi-a-GCST90018940) | 40 | 0.156 | 0.093 | 0.095 | 1.169 | (0.973, 1.404) |
| Hypertension<br>(ebi-a-GCST90038604) | Ventricular arrhythmia<br>(ebi-a-GCST90018940) | 182 | 0.545 | 0.395 | 0.167 | 1.725 | (0.795, 3.742) |
| Heart failure<br>(ebi-a-GCST009541) | Ventricular arrhythmia<br>(ebi-a-GCST90018940) | 5 | 0.403 | 0.193 | 0.036 | 1.497 | (1.026, 2.184) |

### Two SNPs link sleep apnoea and atrioventricular block

The MVMR analysis identified two single nucleotide polymorphisms (SNPs), rs9937053 and rs10507084, as being associated with the causal relationship between sleep apnoea and atrioventricular block, as shown in Figure 1. SNP rs9937053 has been linked to various conditions, including sleep apnoea, obesity, body mass index (BMI), coffee consumption, antithrombotic medication use, and body fat percentage. Univariate Mendelian randomization analysis indicates that body fat percentage, BMI, and obesity have a causal effect on atrioventricular block, whereas coffee consumption and antithrombotic medication use do not (Supplementary Table 2). Furthermore, the MR analysis revealed a bidirectional causal relationship between sleep apnoea and both obesity and BMI, while a unidirectional causal relationship exists between body fat percentage and sleep apnoea (Supplementary Table 3).

**Fig. 1.**
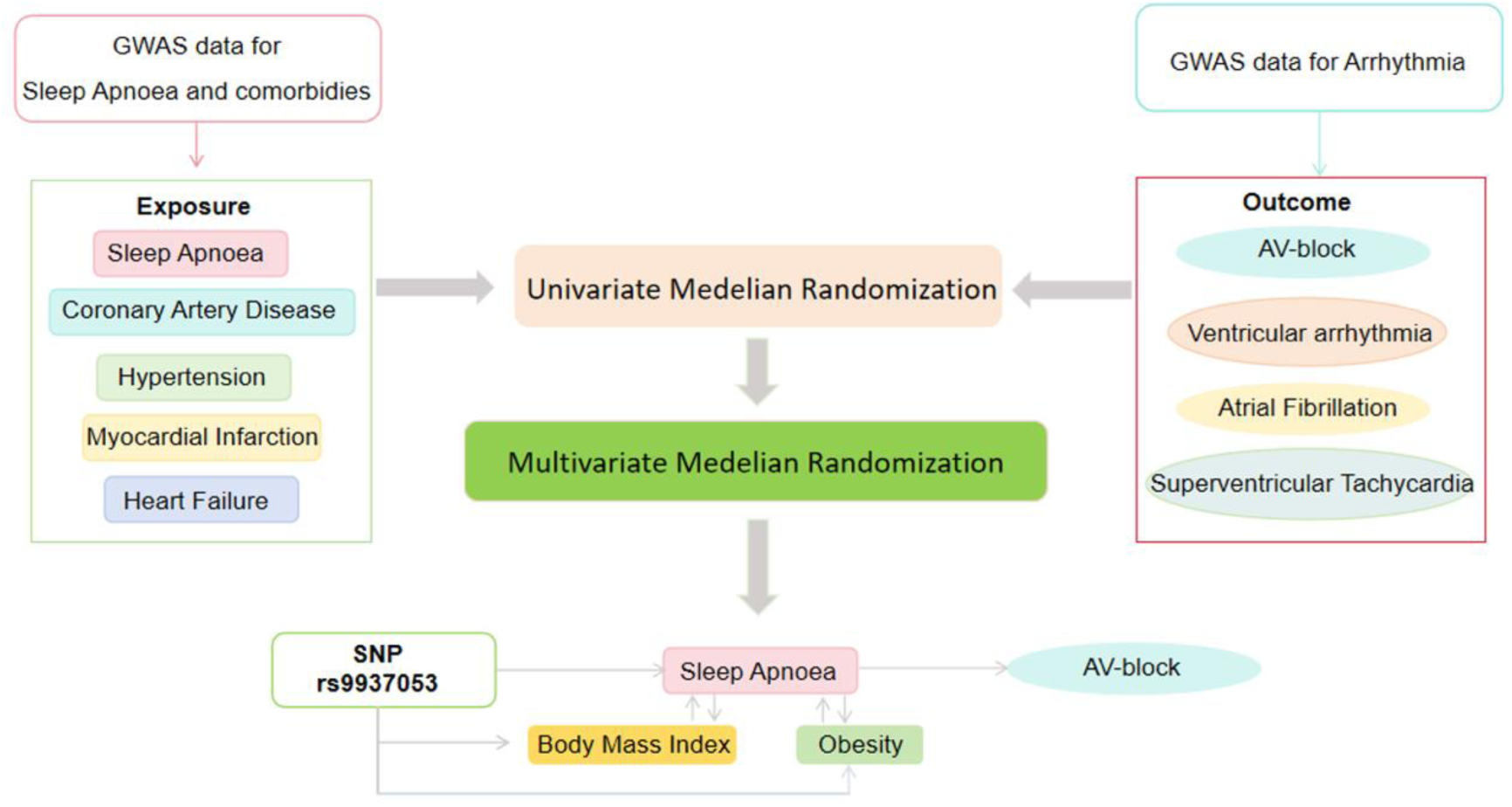
Study diagram and major results.

### Sensitivity analyses

The MR-Egger intercept analysis showed no evidence of horizontal pleiotropy across all MR results (Table 3). However, heterogeneity was observed in the two-sample Mendelian randomization analyses for coronary heart disease, hypertension, heart failure, and atrial fibrillation (Supplementary Table 4). In contrast, the other two-sample Mendelian randomization analyses showed no signs of heterogeneity. This indicates that the results of the two-sample Mendelian randomization analysis assessing the relationship between sleep apnoea exposure and the outcome of atrioventricular block do not exhibit heterogeneity.

**Table 3.** No existence of horizontal pleiotropy between exposure and outcome based on the MR-Egger intercept analysis.

| Exposure (ID) | Outcome (ID) | Egger_intercept | SE | P-value |
| --- | --- | --- | --- | --- |
| Sleep apnoea<br>(finn-b-G6_SLEEPAPNO) | AV-block<br>(finn-b-I9_AVBLOCK) | 0.045 | 0.071 | 0.570 |
| Hypertension<br>(ebi-a-GCST90038604) | AV-block<br>(finn-b-I9_AVBLOCK) | -0.003 | 0.007 | 0.624 |
| Sleep apnoea<br>(finn-b-G6_SLEEPAPNO) | Atrial fibrillation<br>(ebi-a-GCST006414) | -0.018 | 0.025 | 0.520 |
| Coronary artery disease | Atrial fibrillation | -4.78446E-05 | 0.003 | 0.986 |
| (ebi-a-GCST005195) | (ebi-a-GCST006414) |  |  |  |
| Hypertension | Atrial fibrillation | 0.002 | 0.002 | 0.375 |
| (ebi-a-GCST90038604) | (ebi-a-GCST006414) |  |  |  |
| Heart failure | Atrial fibrillation | -0.139 | 0.097 | 0.201 |
| (ebi-a-GCST009541) | (ebi-a-GCST006414) |  |  |  |
| Myocardial infarction | Atrial fibrillation | -0.016 | 0.011 | 0.159 |
| (finn-b-I9_MI_STRICT) | (ebi-a-GCST006414) |  |  |  |
| Sleep apnoea | Ventricular arrhythmia | 0.020 | 0.088 | 0.837 |
| (finn-b-G6_SLEEPAPNO) | (ebi-a-GCST90018940) |  |  |  |
| Coronary artery disease | Ventricular arrhythmia | 0.006 | 0.010 | 0.515 |
| (ebi-a-GCST005195) | (ebi-a-GCST90018940) |  |  |  |
| Hypertension | Ventricular arrhythmia | 0.003 | 0.006 | 0.677 |
| (ebi-a-GCST90038604) | (ebi-a-GCST90018940) |  |  |  |
| Heart failure | Ventricular arrhythmia | 0.006 | 0.038 | 0.884 |
| (ebi-a-GCST009541) | (ebi-a-GCST90018940) |  |  |  |

### Arrhythmia Impact on Sleep Apnoea: Reverse MR Analysis

Despite the univariate Mendelian randomization analysis indicating a causal relationship between sleep apnoea and atrioventricular block, atrial fibrillation, and ventricular arrhythmias, there is no evidence to suggest the presence of reverse causality (Table 4).

**Table 4.**
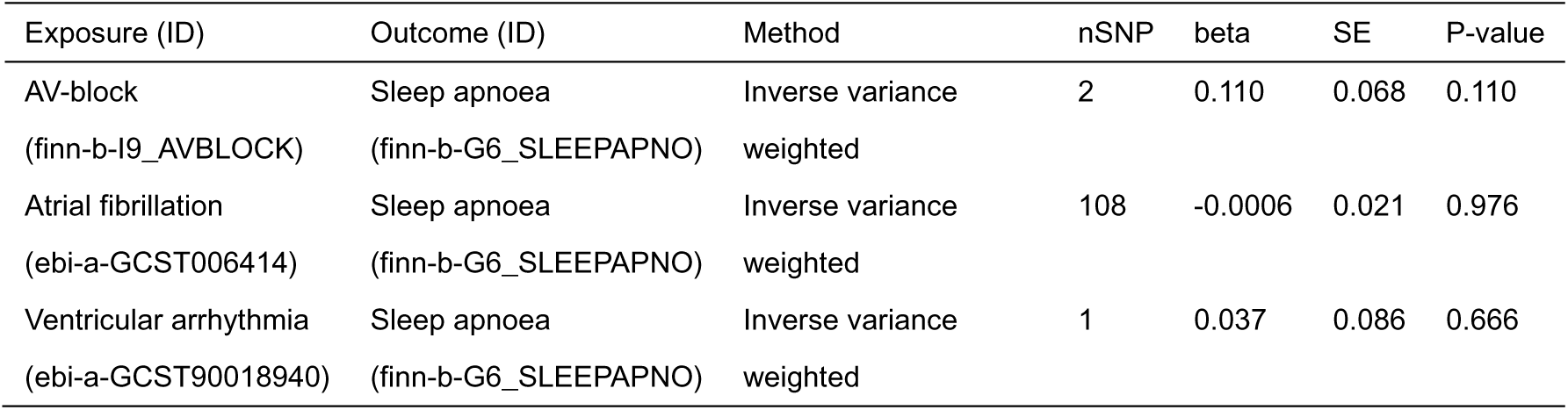
Effects of arrhythmia on sleep apnoea in reverse Mendelian randomization analysis.

## Discussion

The multivariate MR analysis found no correlation between sleep apnoea and ventricular arrhythmias or atrial fibrillation in patients with existing cardiovascular diseases. However, we did identify an independent causal relationship between sleep apnoea and atrioventricular block. Sleep apnoea exhibited a bidirectional causal relationship with hypertension, with both conditions potentially influencing each other and possibly leading to atrioventricular block. Additionally, we identified two SNPs, rs9937053 and rs10507084, that impact the relationship between sleep apnoea and atrioventricular block. Notably, rs9937053 is also associated with body mass index, body fat percentage, and obesity. Further research revealed a bidirectional causal relationship between sleep apnoea and both body mass index and obesity.

Sleep apnoea is associated with low oxygen levels, disruptions in the autonomic nervous system, and alterations in cardiac structure, all of which elevate the risk of developing arrhythmias. Numerous prior investigations have indicated a significant overlap in risk factors between sleep apnoea and atrial fibrillation, with both conditions frequently coexisting^14^. The prevalence of sleep apnoea in patients with AF ranges from 21% to 74%^15–22^. Observational studies suggest that continuous positive airway pressure (CPAP) therapy for OSA could aid in maintaining sinus rhythm post-electrical cardioversion and enhance success rates of catheter ablation^16^. For instance, the SLEEP-AF study demonstrated CPAP therapy’s potential to reverse atrial remodeling in individuals with AF, underscoring the benefits of OSA treatment^23^. Screening for OSA in AF patients is recommended according to the 2020 ESC guidelines^24^. The HARMS2-AF score, which includes a sleep apnoea component, has been proposed to identify individuals at risk of AF development within the general population^25^. However, the causal relationship between sleep apnoea and AF remains ambiguous. While our univariable MR analysis suggested a causal link between sleep apnoea and AF, multivariable MR analysis contradicted this, indicating no causal association. Instead, only heart failure and myocardial infarction exhibited independent causal associations with atrial fibrillation.

Epidemiological studies have shown that patients with sleep apnoea are twice as likely to experience non sustained ventricular tachycardia as normal individuals, and the risk of developing complex ventricular ectopia increases by 50%^26^. In the context of heart failure, both OSA and CSA have been identified as predictors of sleep-related ventricular tachyarrhythmias with fatal outcomes. Elevated central sleep apnoea levels, lasting over 20% of recording time, correlate with increased ventricular tachyarrhythmia incidence, especially during sleep, in heart failure patients with reduced ejection fraction^27^. Limited clinical trials suggest a reduction in ventricular tachyarrhythmia burden through OSA treatment, particularly in heart failure patients^28^. Our univariate MR analysis findings highlight a significant unidirectional causal relationship between coronary artery disease, hypertension, heart failure, and sleep apnoea with ventricular arrhythmias. However, rigorous multivariate MR analysis identified only heart failure retaining a causal association with ventricular arrhythmias.

Second-degree atrioventricular block and higher degrees of heart block during sleep are observed in approximately 4.1% to 16.1% of individuals with sleep apnoea^29–31^. A significantly high prevalence of sleep apnoea syndrome (59%) is noted in patients with long-term pacing^32^. Some reports suggest resolution of atrioventricular block in patients with OSA following CPAP therapy^33,34^. For example, in a study, 651 episodes of heart block were recorded during sleep in sixteen patients with sleep apnoea, with total arrhythmia frequency reduced from 651 to 72 episodes (p<0.01) following nasal CPAP/nasal bilevel positive airway pressure (nBiPAP) therapy^34^. Our multivariate MR analysis identified an independent causal relationship between sleep apnoea and atrioventricular block. However, the mechanism underlying atrioventricular block occurrence in sleep apnoea patients remains unclear. Heart block was completely reversible with atropine administration in patients with no conduction system abnormalities, supporting the concept of elevated vagal tone as a contributing factor^35,36^. Rapid eye movement sleep was identified as a significant independent factor contributing to heart block, suggesting abnormal parasympathetic responsiveness as an additional contributing factor^30^. Additionally, increased systemic blood pressure may affect Atrial-Ventribular conduction through baroreflex mechanisms in sleep apnoea^37^.

Chronic sleep apnoea demonstrates a significant association with arterial hypertension, as evidenced by extensive cross-sectional, longitudinal studies, and a prospective study accounting for potential confounders and demonstrating a dose-response pattern independent of age, BMI, gender, alcohol consumption, and cigarette use^38–41^. Recent post hoc analyses suggest a potential link between moderate to severe OSA and an increased risk of developing more severe forms of hypertension in middle-aged men following a 7.5-year follow-up^42^. Another study found moderate to severe OSA prevalent in 26% of blacks with hypertension, suggesting a link to resistant hypertension^43^. The mechanism involves intermittent hypoxia-reoxygenation, hemodynamic imbalances, and frequent awakenings, leading to decreased nitric oxide levels, elevated catecholamine levels, and increased blood pressure^44^.

Our results identified rs9937053 in the causal relationship between sleep apnoea and atrioventricular block, as determined by MVMR analysis. This SNP is associated with sleep apnoea, obesity, body mass index, and body fat percentage. Our MR analysis also revealed a bidirectional causal relationship between obesity/body mass index and sleep apnoea (Supplementary Table 5). Obesity contributes to sleep apnoea by altering upper airway structure and reducing functional residual capacity, disrupting the respiratory drive-load compensation balance^45^. Central obesity, associated with metabolic abnormalities, strongly correlates with sleep apnoea severity. Furthermore, sleep apnoea’s effects on sleep duration and quality can impact appetite-regulating hormones and metabolism, potentially contributing to weight gain^46^. Thus, sleep apnoea and obesity likely intertwine through bidirectional pathways.

## Acknowledgements

We acknowledge the participants and investigators of the FinnGen study.

## Conflict of interest

The authors declare no competing interests.

## Author contributions

Conception and design: DL and LN; provision of study materials or patients: D.W.W, and M.C; collection and assembly of data: WH, JW, JN, DWW; data analysis and interpretation: WH, JW, and DL; manuscript writing: WH, DL, and LN; final approval of manuscript: all authors; study supervision: DWW and LN; accountable for all aspects of the work: DL and LN. All authors were responsible for reviewing and revising the manuscript.

## Funding

The author(s) disclosed receipt of the following financial support for the research, authorship, and/or publication of this article. This work was supported in part by the projects of National Natural Science Foundation of China (Nos. 82070354 and 81470519) and Program for Huazhong University of Science and Technology Academic Frontier Youth Team, 2019QYTD08 (LN).

## Data Availability

The data that support this work was available in the IEU OpenGWAS Project (https://gwas.mrcieu.ac.uk/), and FinnGen consortium (https://finngen.fi/en), last accessed on 4 July 2023. All data are available in the main text or the supplementary materials.

## Notes

### Competing Interest Statement

The authors have declared no competing interest.

